# Sensor Estimated Home 6-Minute Walk Distance and Cardiac Effort in Pulmonary Hypertension and Heart Failure

**DOI:** 10.1101/2025.09.18.25336075

**Authors:** Daniel Lachant, Nishit Agarwal, Kyle Norton, Deborah Haight, Sebastian Alphonse, Brett Meyer, Kevin Machado Gamboa, Paolo Depetrillo, Hailey Patterson, Becca Slepian, Marvin Slepian, R James White, Melissa Ceruolo

## Abstract

**Background:** Sensor-based estimation of six-minute walk distance and Cardiac Effort, number of heartbeats during the walk divided by distance walked, can be measured in the home. This study aimed to evaluate 1) the impact of walking course length on walk distance and Cardiac Effort, and 2) assess changes over time in a cohort of pulmonary hypertension and heart failure participants.

**Methods:** This was a prospective observational study conducted at two sites. Participants included pulmonary hypertension, heart failure, and healthy controls. All participants wore chest-based sensors capable of recording accelerometry and electrocardiographic data. At baseline visit, participants performed a six-minute walk test on both a 90-ft and a 30-ft course, separated by 30-minutes rest. Unsupervised home walk tests were performed on the following two days. All assessments were repeated eight weeks later. Walk distance and Cardiac Effort were measured using three methods: direct observation, lap count estimation, and a signal-derived method based on mean amplitude deviation. Comparisons between clinic and home-based measurements were made using paired t-tests, correlation analysis, and Bland-Altman testing.

**Results:** Sixty-six participants were enrolled, including 42 with pulmonary hypertension or heart failure and 24 controls. Walk distance on the 90-foot course was 10.7 percent longer than on the 30-ft course. Home-based walk distance was similar to the 30-ft course but lower than the 90-ft course. Home Cardiac Effort was similar to in-clinic values and tracked with changes over time.

**Conclusion:** Home-based functional testing incorporating Cardiac Effort may offer a simple and effective remote metric to improve management of patients with heart failure.

## Introduction

The 6-minute walk test (6MWT), performed according to American Thoracic Society guidelines, is a simple assessment of functional capacity in a supervised clinical setting(1). It requires minimal equipment, a 30 m unobstructed hallway and a healthcare personnel to count laps to calculate a 6-minute walk distance (6MWD). In pulmonary arterial hypertension (PAH), 6MWD provides insight into disease status and treatment response(2). Unfortunately, current 6MWT is limited on how frequent it can be performed. Cardiac Effort (CE)(3–6), the number of heart beats during the 6MWT divided by 6MWD, was developed to provide deeper insight into the physiologic effort required to achieve a certain 6MWD. It helps differentiate whether changes in 6MWD are due to variations in effort or underlying changes in physiology. CE has been incorporated as an exploratory end point in a PAH clinical trial(7), in pulmonary hypertension with interstitial lung disease (NCT06129240), and in a pulmonary embolism(8) and PAH rehabilitation study (NCT06477640).

The COVID-19 pandemic disrupted routine care in PAH. Interpreting changes in 6MWD became more difficult with masking(9). This led to home 6MWT being performed in pulmonary hypertension (6, 10, 11). All three studies found it was safe to perform 6MWT in the home setting. The first report in PAH found 6MWD at home and in the clinic were similar when performed on 90ft walking space and supervised (10). The second report in PAH showed unsupervised 6MWT performed on a modified walking space (~40ft) using sensor estimated 6MWD was lower than clinic 6MWD, but CE provided a similar measurement (10).The third study in a mix pulmonary hypertension cohort used an app to perform tele-6MWT in a mix group of pulmonary hypertension patients and found tele-6MWT had a bias of 25 m over directly observed 6MWT (11). They did not report how the app performed on directly observed 6MWT.

In this study, we performed in clinic and home 6MWT and CE in a mix pulmonary hypertension and heart failure cohort and evaluated changes overtime. We hypothesized that CE would have less variability when comparing in clinic to home measurements by accounting for changes in heart rate. We did the following, 1) Compared 6MWD and CE on 90-ft and 30-ft walking spaces in the clinic to evaluate the impact of walking space length and the frequency of turns on test results. 2) Compared directly observed, lap-estimated, and mean amplitude deviation (MAD)-estimated 6MWD and CE values obtained in the clinic to those obtained during home monitoring at each time point. 3) Compared changes in 6MWD and CE from baseline to Week-8 during home monitoring to corresponding changes observed in the clinic.

## Methods

This was a prospective observational study conducted at the University of Rochester Medical Center and the University of Arizona. Institutional Review Board approval was obtained. The study development and analysis were performed collaboratively by the authors, with Metadata sponsoring the study. We selected a chest-based form factor for the wearable sensor to ensure the highest data quality, as this placement facilitates superior cardiac data collection via ECG during ambulation compared to wrist-based photoplethysmography (PPG) sensors, which is crucial for accurately computing the Cardiac Effort metric from both heart rate and acceleration data. All participants provided informed consent before any study activities were performed. Participants with treated pulmonary hypertension or heart failure were eligible to participate. We included a small cohort of Heart Failure (HF) patients alongside the Primary Pulmonary Hypertension (PH) cohort to explore the utility of the Cardiac Effort metric in a heterogeneous group of cardiopulmonary-limited patients. There was also a control cohort that was not age sex matched to see the impact of longer walkers. Recruitment occurred between September 2022-February 2024 as they were seen during routine clinic visits. Participants needed to be functional class I-III, ambulatory, and have an unobstructed walking space of at least 30ft at home. Participants did not wear a mask during the in-clinic or at-home study procedures. Safety measures for the at-home walks included requiring a caregiver to be present, with no adverse events reported.

Baseline testing included demographics, clinical characteristics, quality of life questionnaire (emPHasis-10(12) or Minnesota Living with Heart Failure questionnaire(13)), NT-pro BNP, and Reveal lite 2 score(14) in pulmonary hypertension only. The BioStamp nPoint® (Medidata), a medical-grade wearable patch has been previously validated to measure accelerometry and electrocardiography(15) and used in this pilot study(6), was placed on the left upper chest and thigh with disposable adhesive prior to the 6MWT [Figure 1]. Accelerometry and electrocardiography data was recorded at 31.25 Hz and 250 Hz, respectfully. Concurrently we utilized the VitalPatch® (VitalConnect), a disposable wearable patch also measuring accelerometry (50 Hz) and electrocardiography (125 Hz), validated for remote vitals data collection(16, 17). Both devices are FDA cleared for medical use. Participants rested for 3 minutes before the first 6MWT. At each visit participants completed two 6MWT, 90ft walking space (1) and a second on a 30ft walking space, with a 30 minute rest between 6MWT. After the second 6MWT, participants went home wearing one of the devices. Due to supply chain issues, 26 subjects wore BioStamp nPoint only for the two visits, 24 wore both BioStamp nPoint for two visits and VitalPatch for the second visit only, and 16 wore BioStamp nPoint and VitalPatch during both visits. Data from both devices was collected and transmitted to a secure cloud for analysis. The goal was not to compare sensors. On day 1 and 2, a home 6MWT on a modified walking space was performed. The course length was recorded by the participant using a 100 ft tape measure. Safety assessment was performed to evaluate for syncope, falls, or other safety concerns related to home 6MWT or devices. Eight weeks (+/-7 days) later participants would complete the same evaluation as baseline, only this time the order of 6MWT on the 90ft and 30ft were switched.

**Figure 1.**
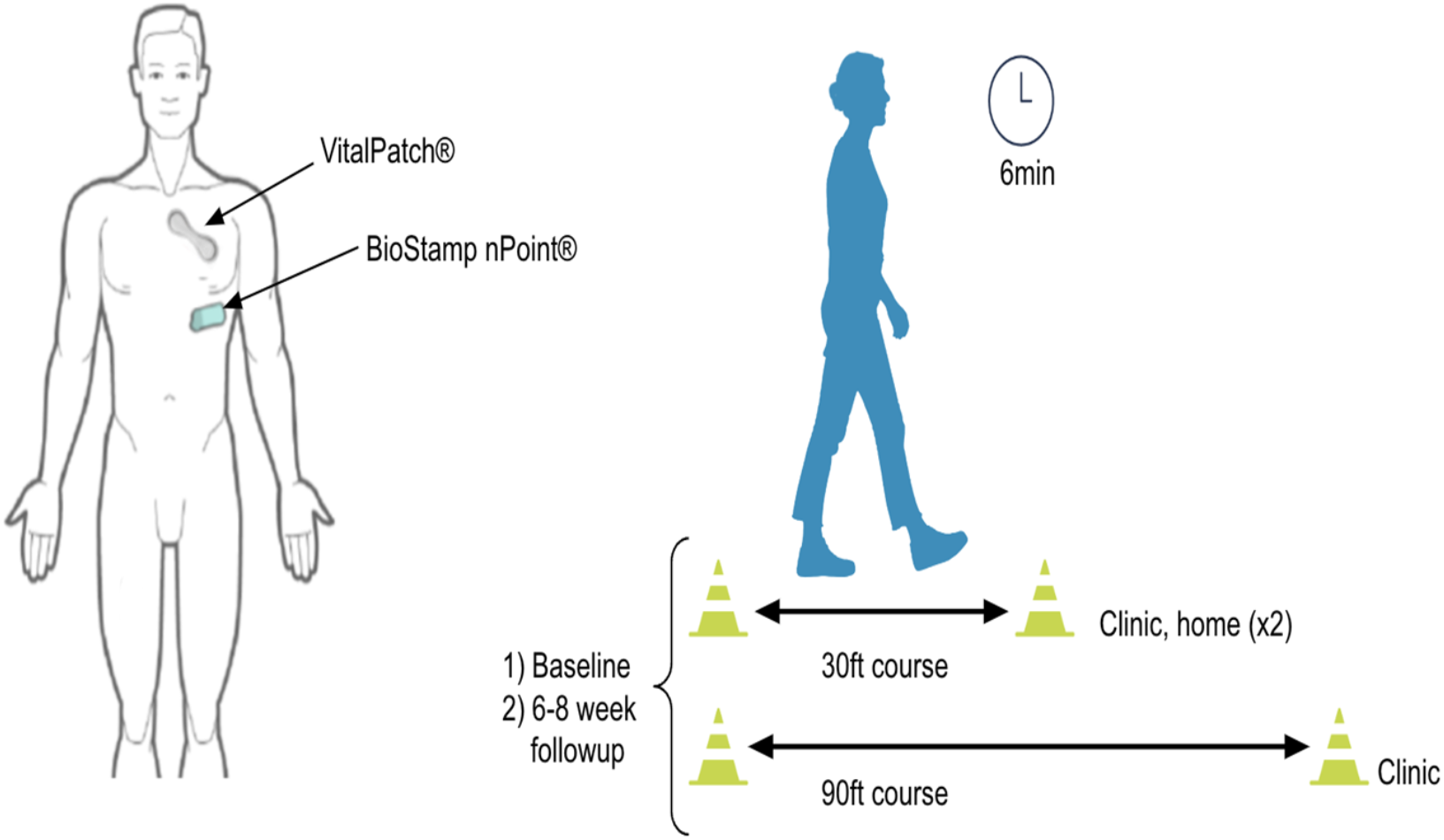
Device placement and study protocol. (Left) Location of BioStamp nPoint and VitalPatch placed concurrently on the chest. (Right) Study protocol: two visits were conducted, each consisting of two 6-minute walk tests (6MWT), one on a 90 ft walking space and the other on a 30 ft walking space. At each visit, participants completed their study protocol and then went home wearing one of the devices.

6MWD in clinic was calculated three different ways on the 90 and 30 ft walking spaces: 1) directly observed distance (6MWD_D_), 2) estimating distance based on lap counting with accelerometry (6MWD_L_, as previously described)(6), and 3) estimating distance based on changes in mean amplitude deviation (MAD) (6MWD_M,_ as previously described, distance estimated by changes in acceleration units)(6). These were the same methods used in this prior report(6). In the home setting, 6MWD was estimated using 1) lap counting and 2) changes in MAD. Cardiac Effort (CE) was calculated as previously described(4, 6) using all three 6MWD methods (CE_D_, CE_L_, CE_M_). The electrocardiography heart rate data was analyzed using LabChart 8 (AdInstruments).

Categorical variables were reported as counts and percent. Continuous variables were reported as median with interquartile range. Correlations were performed with Pearson or Spearman depending on the distribution of the data. Bland-Altman was used to compare measurements. We compared directly observed 6MWD and Cardiac Effort to estimated methods. The 90ft measurements were compared to 30ft measurements. The in-clinic measurements were compared to home measurements and repeat in clinic and home measurements were compared for reproducibility. Parametric and non-parametric t testing were performed depending on the distribution of the data.

## Results

### Baseline demographics

Sixty-six participants were enrolled, 31 with pulmonary hypertension, 11 heart failure, and 24 controls [Table 1]. Th majority of pulmonary hypertension participants were middle aged, the heart failure participants were older, and the healthy controls were younger. There were 10 participants with functional class III symptoms. The baseline 6MWD_D_ and CE_D_ for all participants were 434 m and 1.47 beats/m [Table 1]. The average home walking space was 37ft. Out of 459 walks, 26 electrocardiography tracings were not usable because of artifacts (Supplement Table 1]). All 66 participants completed baseline testing. However, the longitudinal analysis comparing baseline clinic data against the eight-week home follow-up data was limited to 55 participants. The remaining 11 participants (5 healthy controls, 4 PAH, and 2 heart failure patients) were excluded from the longitudinal cohort as they only had data available from the baseline visit. Clinical worsening contributed to non-control participants missing the week-8 visit. For controls, relocating and COVID infections limited week-8 follow up. There was near 100% compliance with walks at home with only two home walks not being performed.

**Table 1.**
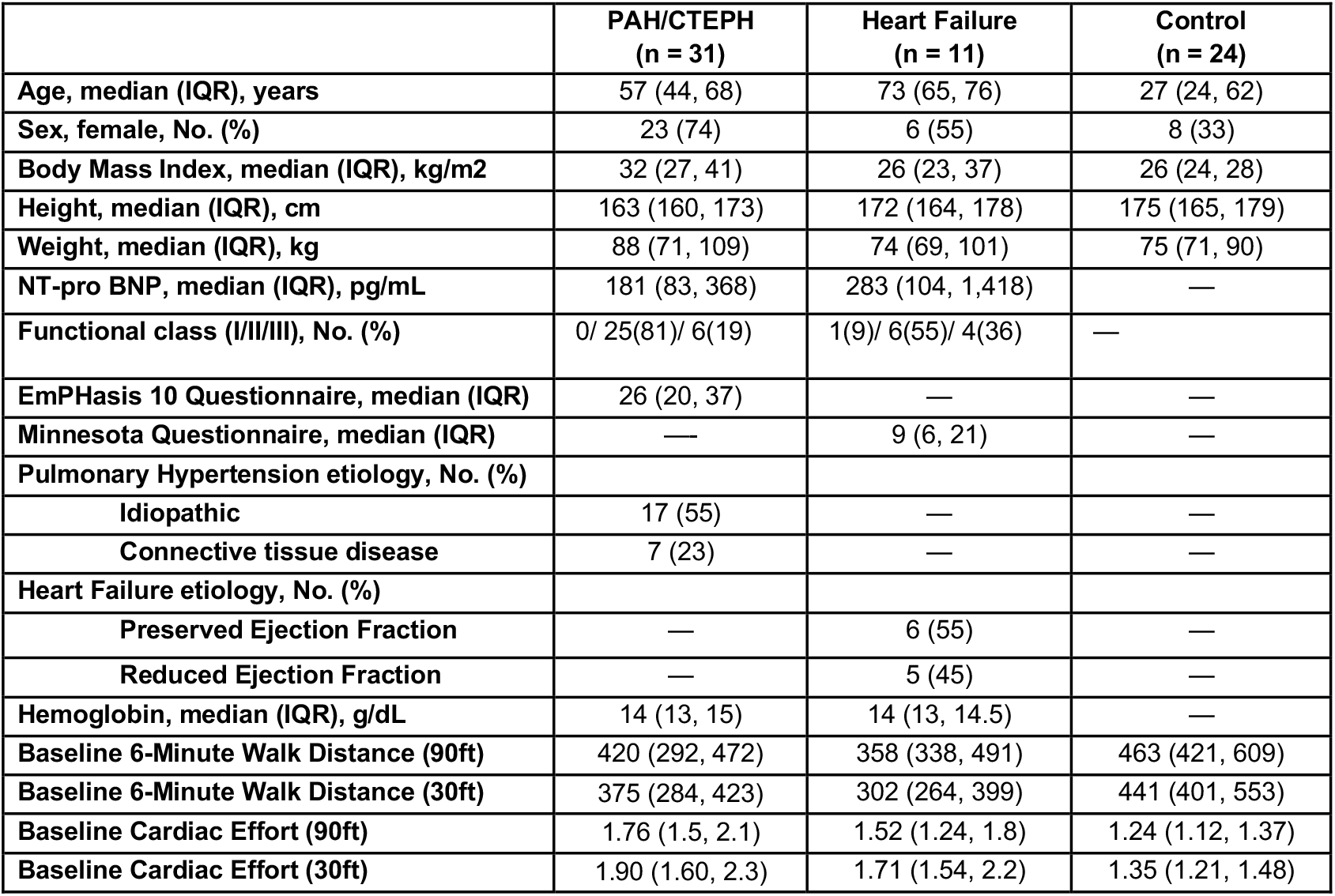
Baseline Demographics.

|  | **PAH/CTEPH**<br>**(n = 31)** | **Heart Failure**<br>**(n = 11)** | **Control**<br>**(n = 24)** |
| --- | --- | --- | --- |
| **Age, median (IQR), years** | 57 (44, 68) | 73 (65, 76) | 27 (24, 62) |
| **Sex, female, No. (%)** | 23 (74) | 6 (55) | 8 (33) |
| **Body Mass Index, median (IQR), kg/m<sup>2</sup>** | 32 (27, 41) | 26 (23, 37) | 26 (24, 28) |
| **Height, median (IQR), cm** | 163 (160, 173) | 172 (164, 178) | 175 (165, 179) |
| **Weight, median (IQR), kg** | 88 (71, 109) | 74 (69, 101) | 75 (71, 90) |
| **NT-pro BNP, median (IQR), pg/mL** | 181 (83, 368) | 283 (104, 1,418) | — |
| **Functional class (I/II/III), No. (%)** | 0/ 25(81)/ 6(19) | 1(9)/ 6(55)/ 4(36) | — |
| **EmPHasis 10 Questionnaire, median (IQR)** | 26 (20, 37) | — | — |
| **Minnesota Questionnaire, median (IQR)** | — | 9 (6, 21) | — |
| **Pulmonary Hypertension etiology, No. (%)** |  |  |  |
| **Idiopathic** | 17 (55) | — | — |
| **Connective tissue disease** | 7 (23) | — | — |
| **Heart Failure etiology, No. (%)** |  |  |  |
| **Preserved Ejection Fraction** | — | 6 (55) | — |
| **Reduced Ejection Fraction** | — | 5 (45) | — |
| **Hemoglobin, median (IQR), g/dL** | 14 (13, 15) | 14 (13, 14.5) | — |
| **Baseline 6-Minute Walk Distance (90ft)** | 420 (292, 472) | 358 (338, 491) | 463 (421, 609) |
| **Baseline 6-Minute Walk Distance (30ft)** | 375 (284, 423) | 302 (264, 399) | 441 (401, 553) |
| **Baseline Cardiac Effort (90ft)** | 1.76 (1.5, 2.1) | 1.52 (1.24, 1.8) | 1.24 (1.12, 1.37) |
| **Baseline Cardiac Effort (30ft)** | 1.90 (1.60, 2.3) | 1.71 (1.54, 2.2) | 1.35 (1.21, 1.48) |

### Safety

Importantly, with 10 functional class III and 5 participants on supplemental oxygen, there were no issues related to the devices, falls, tripping on oxygen tubing, syncope, or unexpxected emergency room visits related to the home 6MWT.

### 6MWD and Cardiac Effort 90 and 30 ft In Clinic

6MWD_D_ and CE_D_ on 90 and 30 ft for all participants are listed in [Table 2]. The 6MWD_D_ and CE_D_ were both better on the 90ft walking space; even when 30ft walking course was performed first. Positive correlations were observed between measurements obtained on 90ft and 30ft walking spaces for both 6MWD_D_ (r=0.966, p<0.005) and CE_D_ (r=0.941, p<0.005), respectively. In Figure 2, Bland-Altman analysis revealed mean differences of 10.73% for 6MWD_D_ and −9.91% for CE_D_ between measurements obtained on 90ft and 30ft walking spaces.

**Table 2.** 6-Minute Walk Distance and Cardiac Effort at Baseline and Week 8.

|  | 6-Minute Walk Distance |  |  |  |  |  |
| --- | --- | --- | --- | --- | --- | --- |
|  | Directly Observed |  | Lap Estimated |  | Mean Amplitude Deviation |  |
|  | PH/HF<br>(n = 42) | Control<br>(n = 24) | PH/HF<br>(n = 42) | Control<br>(n = 24) | PH/HF<br>(n = 42) | Control<br>(n = 24) |
| **Baseline 6MWD (90ft) – Clinic, (IQR), m** | 416<br>(317,472) | 463<br>(421, 609) | 384<br>(285, 466) | 452<br>(411, 604) | 404<br>(315, 483) | 486<br>(420, 575) |
| **Baseline 6MWD (30ft) – Clinic, (IQR), m** | 373<br>(284, 416) | 441<br>(401, 553) | 370<br>(269, 413) | 430<br>(388, 550) | 370<br>(282, 439) | 461<br>(404, 551) |
| **Baseline Cardiac Effort (90 ft) – Clinic, (IQR), beats/m** | 1.7<br>(1.5, 2.2) | 1.2<br>(1.1, 1.4) | 1.7<br>(1.5, 2.2) | 1.3<br>(1.1,1.4) | 1.7<br>(1.5, 2.0) | 1.3 (1.1, 1.4) |
| **Baseline Cardiac Effort (30 ft) – Clinic, (IQR), beats/m** | 1.8<br>(1.6, 2.4) | 1.4<br>(1.2, 1.5) | 1.9<br>(1.6, 2.3) | 1.4<br>(1.3, 1.5) | 1.8<br>(1.6, 2.1) | 1.3 (1.2, 1.4) |
| **Baseline 6MWD – Home, (IQR), m** | - | - | 373<br>(305, 436) | 502<br>(395, 579) | 375<br>(287, 449) | 493<br>(405, 574) |
| **Baseline Cardiac Effort – Home, (IQR), beats/m** | - | - | 1.8<br>(1.5, 2.2) | 1.4<br>(1.3, 1.5) | 1.9<br>(1.6, 2.1) | 1.4<br>(1.3, 1.5) |
| **Week 8 6MWD (90ft) – Clinic, (IQR), m** | 429<br>(305, 471) | 529<br>(463, 615) | 419<br>(304, 470) | 519<br>(418, 616) | 434<br>(336, 482) | 538<br>(483, 576) |
| **Week 8 6MWD (30ft) – Clinic, (IQR), m** | 380<br>(283, 419) | 469<br>(404, 564) | 391<br>(285, 427) | 467<br>(398, 563) | 379<br>(317, 441) | 501<br>(458, 539) |
| **Week 8 Cardiac Effort (90 ft) – Clinic, (IQR), beats/m** | 1.7<br>(1.5, 2.1) | 1.1<br>(1.1, 1.3) | 1.7<br>(1.4, 2.2) | 1.2<br>(1.1, 1.4) | 1.6<br>(1.5, 1.9) | 1.2<br>(1.1, 1.3) |
| **Week 8 Cardiac Effort (30 ft) – Clinic, (IQR), beats/m** | 1.9<br>(1.6, 2.3) | 1.4<br>(1.2, 1.5) | 1.8<br>(1.6, 2.3) | 1.4<br>(1.2, 1.5) | 1.7<br>(1.6, 2.1) | 1.3<br>(1.1, 1.3) |
| **Week 8 6MWD – Home, (IQR), m** | - | - | 347<br>(290, 410) | 486<br>(347, 548) | 348<br>(266, 412) | 506<br>(353, 557) |
| **Week 8 Cardiac Effort – Home, (IQR), beats/m** | - | - | 1.8<br>(1.6, 2.4) | 1.4<br>(1.2, 1.6) | 1.8<br>(1.7, 2.2) | 1.3<br>(1.2, 1.5) |

6-Minute Walk Distance (6MWD), Pulmonary Hypertension (PH), Heart Failure (HF), Median with interquartile ranges.

**Figure 2.**
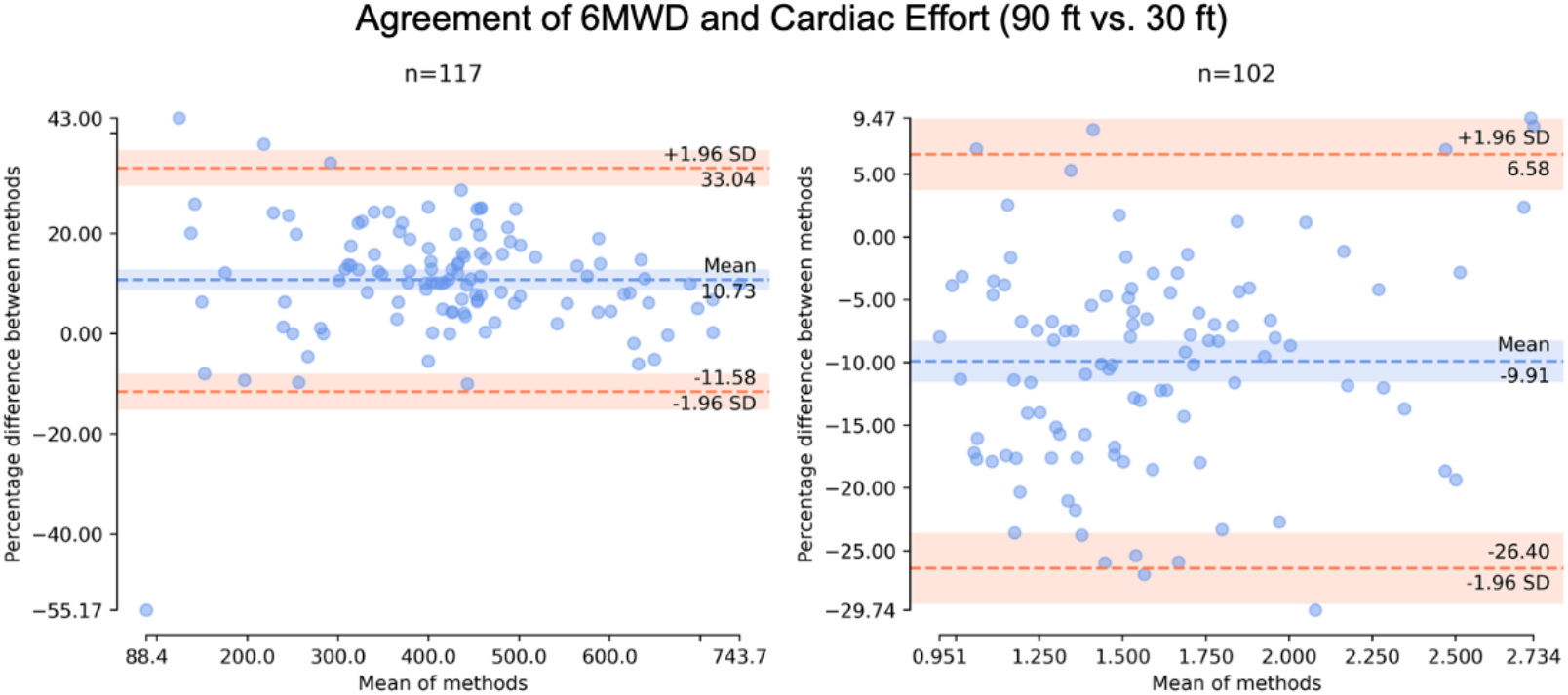
Agreement analysis for 6-minute walk distance (6MWD) and Cardiac Effort (CE) measurements obtained on different walking space lengths. (Left) Bland Altman Plot showing the agreement between 6MWD values obtained on a 90 ft vs 30 ft walking space. (Right) Bland Altman Plot showing the agreement between CE values obtained on a 90 ft vs 30 ft walking space.

### 6MWD and Cardiac Effort Estimate In Clinic

There were no differences between 6MWD_D_ on 90ft walking space or 30ft walking space with 6WMD_L_ (n=118), (447 m (366, 515) vs 429m (356, 506) p=0.22) and (401 m (313, 454) vs 399 m (321, 461) p=0.96). In Figure 3–4, Bland Altman analysis shows that 6WMD_L_ was 3.79% different from 6WMD_D_ on the 90ft walking space and 1.40% on 30 ft walking space. Five walks could not have a distance estimated because turns could not be identified. Correlation analysis shows 6WMD_L_ is positively correlated to 6WMD_D_ on the 90ft walking space (r=0.97, p<0.005) and on the 30 ft walking space (r=0.99, p<0.005).

**Figure 3.**
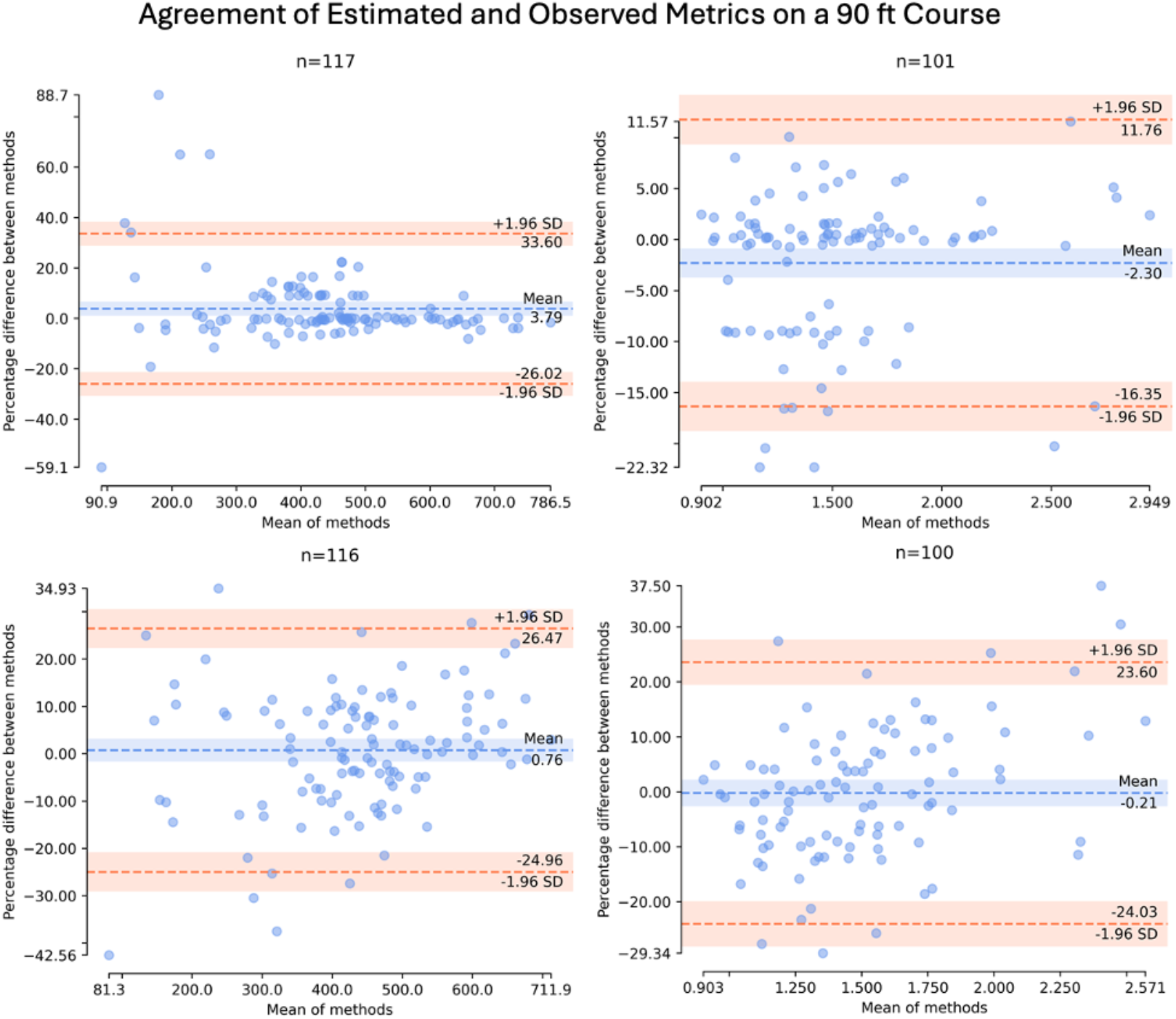
Agreement between estimated and directly observed walk distances and cardiac efforts on a 90ft walking space. (Top left) Bland-Altman plot of lap-estimated 6MWD vs directly observed 6MWD on a 90ft walking space. (Top right) Bland-Altman plot of lap-estimated Cardiac Effort vs directly observed Cardiac Effort on a 90ft walking space. (Bottom left) Bland-Altman plot of mean amplitude deviation estimated 6MWD vs directly observed 6MWD on a 90ft walking space. (Bottom right) Bland-Altman plot of mean amplitude deviation estimated Cardiac Effort vs directly observed Cardiac Effort on a 90ft walking space

**Figure 4.**
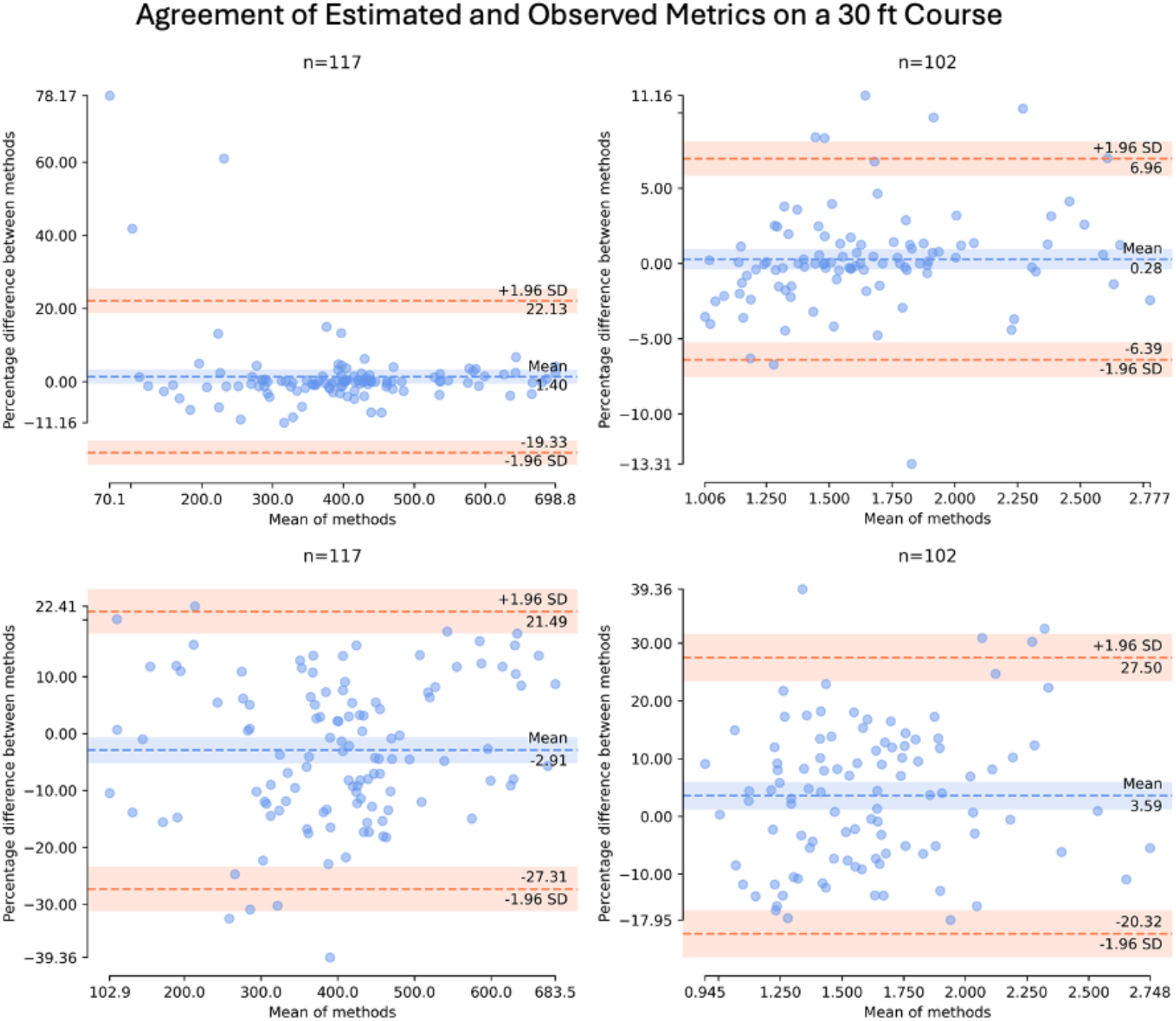
Agreement between estimated and directly observed walk distances and cardiac efforts on a 30ft walking space. (Top left) Bland-Altman plot of lap-estimated 6MWD vs directly observed 6MWD on a 30ft walking space. (Top right) Bland-Altman plot of lap-estimated Cardiac Effort vs directly observed Cardiac Effort on a 30ft walking space. (Bottom left Bland-Altman plot of mean amplitude deviation estimated 6MWD vs directly observed 6MWD on a 30ft walking space. (Bottom right) Bland-Altman plot of mean amplitude deviation estimated Cardiac Effort vs directly observed Cardiac Effort on a 30ft walking space.

There was no difference between 6MWD_D_ on 90ft walking space and 6WMD_M_ (n=117) (445 m (366, 515) vs 441 m (380,514) p=0.21). The 6MWD_D_ on the 30 ft walking space was shorter than the 6MWD_M_ (401 m (313, 454) vs 412 m (338,481), p=0.03). This could be from movement during extra turns that MAD captures. In Figure 3–4, Bland Altman analysis shows that 6MWD_M_ was 0.76% different from 6MWD_D_ on the 90ft walking space and −2.91% on 30 ft walking space. Correlation analysis shows 6WMD_M_ is positively correlated to 6WMD_D_ on the 90ft walking space (r=0.93, p<0.005) and on the 30 ft walking space (r=0.93, p<0.005).

There was no difference between CE_D_ on 90ft walking space or 30ft walking space with CE_L_ (n=113), (1.47 beats/m (1.20, 1.81) vs 1.51 beats/m (1.27, 1.81), p=0.36) and (1.62 beats/m (1.35, 1.99) vs 1.60 (1.36, 1.94) p=0.47). In Figure 3–4, Bland Altman analysis shows that CE_L_ was −2.3% different from CE_D_ on the 90ft walking space and 0.28% on 30ft walking space. Correlation analysis shows CE_L_ is highly correlated to CE_D_ on the 90ft walking space (r=0.97, p<0.005) and on the 30 ft walking space (r=0.99, p<0.005).

There was no significant difference between CE_D_ on 90ft walking space and CE_M_ (n=112) (1.47 beats/m (1.20, 1.81) vs 1.49 beats/m (1.24, 1.76) p=0.56). The CE_D_ on the 30 ft walking space was higher than the CE_M_ (1.62 beats/m (1.35, 1.99) vs 1.58 (1.33, 1.88), p=0.02). This could be from acceleration changes related to the extra turns that mean amplitude captures. In Figure 3–4, Bland Altman analysis shows that CE_M_ was −0.21% different from directly observed Cardiac Effort on the 90ft walking space and 3.59% on 30 ft walking space. Correlation analysis shows CE_M_ is highly correlated to CE_D_ on the 90ft walking space (r=0.87, p<0.005) and on the 30 ft walking space (r=0.85, p<0.005).

### In clinic vs at home Lap Estimated 6MWD and Cardiac Effort

Using 6MWD_L_, the 90ft clinic 6MWD_L_ was further than the home 6MWD_L_ (baseline, 434 m (367, 502) vs 400 m (323, 507), p=0.0025) and (week 8, 464 m (368, 566) vs 390 m (329, 508), p<0.0001). There was no difference between 6MWD_L_ on the 30ft walking space with the home 6MWD_L_ (baseline, 400 m (334, 466) vs 400 m (323, 507), p=0.75) and (week 8, 412 m (344, 490) vs 390 m (329, 508), p=0.12). In Figure 5–6, Bland-Altman analysis revealed 6MWD_L_ at home was 8.21% different from 6MWD_L_ in clinic on the 90ft walking space and −0.36% different from 6MWD_L_ in clinic on the 30ft walking space. Correlation analysis revealed 6MWD_L_ at home is correlated to 6MWD_L_ in clinic both on the 90ft (r=0.89, p<0.005) and 30ft (r=0.9, p<0.005) walking spaces.

**Figure 5.**
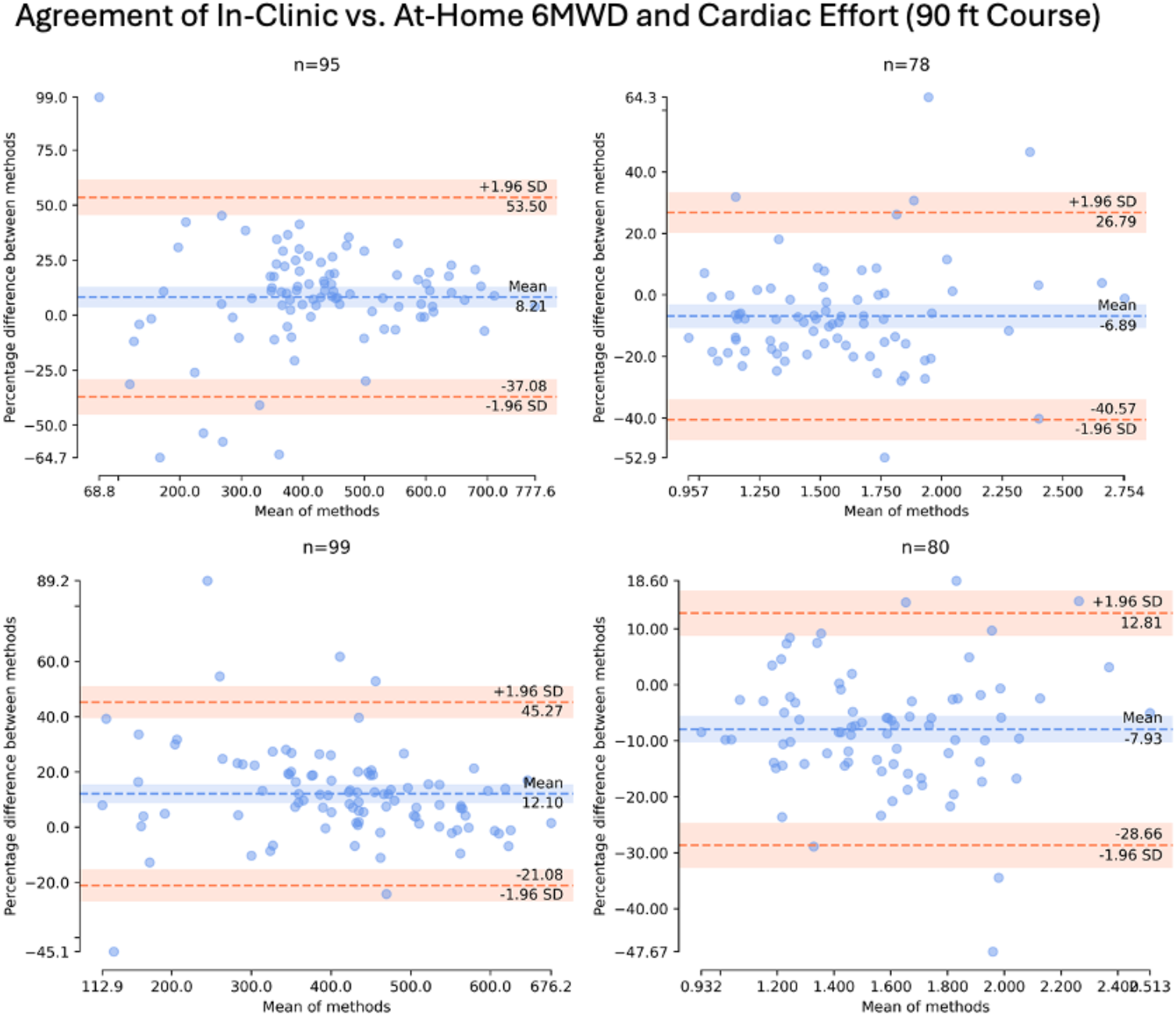
Agreement between in clinic vs at home assessment walk distances and cardiac efforts. (Top left) Bland-Altman plot of lap-estimated 6MWD at home vs lap-estimated 6MWD in clinic on a 90ft walking space. (Top right) Bland-Altman plot of lap-estimated Cardiac Effort at home vs lap-estimated Cardiac Effort in clinic on a 90ft walking space. (Bottom left) Bland-Altman plot of mean amplitude deviation estimated 6MWD at home vs mean amplitude deviation estimated 6MWD in clinic on a 90ft walking space. (Bottom right) Bland-Altman plot of mean amplitude deviation estimated Cardiac Effort at home vs mean amplitude deviation estimated Cardiac Effort in clinic on a 90ft walking space.

**Figure 6.**
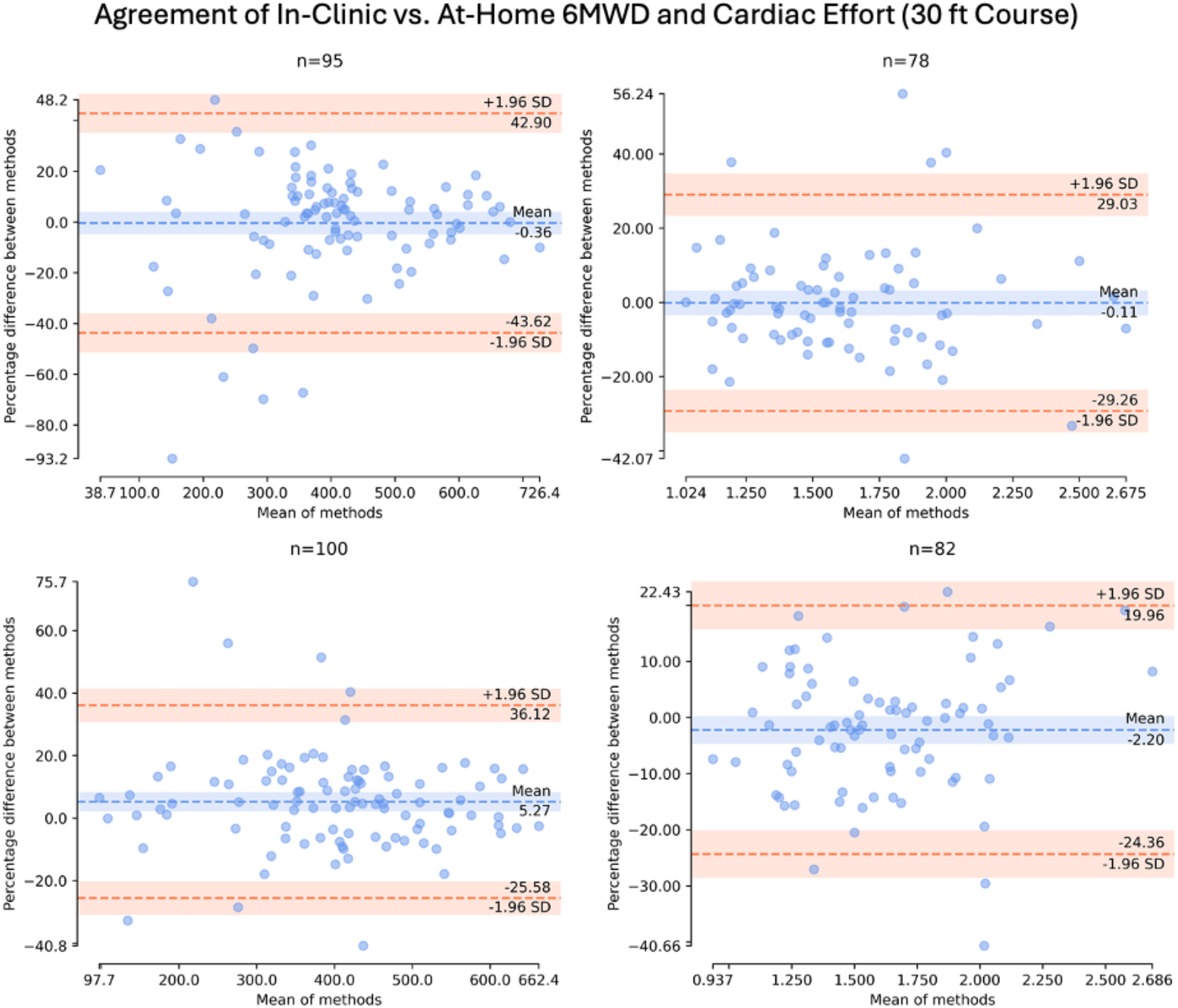
Agreement between in clinic vs at home assessment walk distances and cardiac efforts. (Top left) Bland-Altman plot of lap-estimated 6MWD at home vs lap-estimated 6MWD in clinic on a 30ft walking space. (Top right) Bland-Altman plot of lap-estimated Cardiac Effort at home vs lap-estimated Cardiac Effort in clinic on a 30ft walking space. (Bottom left) Bland-Altman plot of mean amplitude deviation estimated 6MWD at home vs mean amplitude deviation estimated 6MWD in clinic on a 30ft walking space. (Bottom right) Bland-Altman plot of mean amplitude deviation estimated Cardiac Effort at home vs mean amplitude deviation estimated Cardiac Effort in clinic on a 30ft walking space.

Using CE_L_, home CE_L_ was higher than 90ft CE_L_ in clinic at baseline, (1.53 beats/m (1.27, 1.91) vs 1.63 beats/m (1.38, 1.99), p=0.01) and week 8 (1.51 beats/m (1.22, 1.79) vs 1.58 beats/m (1.36, 2.01), p=0.008). There was no difference at baseline or week 8 between CE_L_ on the 30ft walking space with the home CE_L_ (1.58 beats/m (1.36, 2.22) vs 1.63 beats/m (1.39, 1.99), p=0.62) and (1.62 beats/m (1.38, 1.94) vs 1.57 beats/m (1.41, 2), p=0.89). In Figure 5–6, Bland-Altman analysis revealed CE_L_ at home was −6.9% different from CE_L_ in clinic on the 90ft walking space and −0.11% different from CE_L_ in clinic on the 30ft walking space. Correlation analysis revealed CE_L_ at home is correlated to CE_L_ in clinic both on the 90ft (r=0.7, p<0.005) and 30ft (r=0.77, p<0.005) walking spaces.

Using pooled CE_L_ (to account for the different order of in clinic tests completed) at baseline and week 8 from pulmonary hypertension and heart failure participants only, we observed that CE_L_ at home was higher than CE_L_ in clinic for the 90ft walking space in clinic (1.70 beats/m (1.48, 2.24) vs 1.82 beats/m (1.58, 2.26), p=0.03), while 30ft walking space in clinic was similar to at home (1.84 beats/m (1.60, 2.36) vs 1.82 beats/m (1.58, 2.26), p=0.80). Using pooled 6MWD_L_ from pulmonary hypertension and heart failure participants only, we observed 6MWD_L_ in clinic for the 90 ft walking was further than home 6MWD_L_ (415 m (284, 474) vs 365 m (304, 417), p = 0.0003), while there was no difference with 30ft walking space in clinic versus at home (378 m (271, 429) vs 365 m (304,417), p = 0.65).

### In clinic vs at home Mean Amplitude Deviation 6MWD and Cardiac Effort

Using 6MWD_M_, there was a similar finding of the 90ft 6MWD_M_ being further than the home 6MWD_M_ (baseline, 438 m (381,514) vs 411 m (315,490), p<0.0001) and week 8, 449 m (361, 542) vs 390 m (304, 462), p<0.0001). There was no difference at baseline between 6MWD_M_ on the 30ft walking space with the home 6MWD_M_ (419 m (346, 472) vs 411 m (315, 490), p=0.09). However, during week 8 visit we observed a significant difference (415 m (332, 504) vs 392 m (310, 485), p=0.0007). In Figure 5–6, Bland-Altman analysis revealed 6MWD_M_ at home was 12.1% different from 6MWD_M_ in clinic on the 90ft walking space and 5.27% different from 6MWD_M_ in clinic on the 30ft walking space. Correlation analysis revealed 6MWD_M_ at home is correlated to 6MWD_M_ in clinic both on the 90ft (r=0.9, p<0.005) and 30ft (r=0.91, p<0.005) walking spaces.

Using CE_M_, the 90ft CE_M_ was different than what was obtained at home (baseline, 1.51 beats/m (1.35, 1.80) vs 1.67 beats/m (1.40, 2.02), p=0.0001) and week 8, 1.55 beats/m (1.32, 1.85) vs 1.70 beats/m (1.48, 2.0), p=<0.0001). There was no difference at baseline or week 8 between 30 ft CE_M_ with the home CE_M_ (1.57 beats/m (1.39, 1.97) vs 1.67 beats/m (1.40, 2.02), p=0.25) and (1.62 beats/m (1.33, 1.94) vs 1.67 beats/m (1.45, 2.0), p=0.36). In Figure 5–6, Bland-Altman analysis revealed CE_M_ at home was −7.93% different from CE_M_ in clinic on the 90ft walking space and −2.20% different from mean amplitude estimated Cardiac Effort in clinic on the 30ft walking space. Correlation analysis revealed CE_M_ at home is correlated to CE_M_ in clinic both on the 90ft (r=0.85, p<0.005) and 30ft (r=0.85, p<0.005) walking spaces.

Pooled CE_M_ in clinic on 90 ft walking space was different from home CE_M_ (1.69 beats/m (1.51, 2.00) vs 1.84 beats/m (1.65, 2.15), p<0.0001), while there was no difference with 30 ft CE_M_ and home CE_M_ (1.81 beats/m (1.57, 2.13) vs 1.84 beats/m (1.65, 2.15) p=0.60). Pooled 6MWD_M_ in clinic on 90 ft walking space was different from home 6MWD_M_ (413 m (320, 484) vs 348 m (275, 429), p < 0.0001) and 30ft walking space (371 m (295, 446) vs 348 m(275, 429), p = 0.01).

### Changes in clinic vs at home

In pulmonary hypertension and heart failure participants only, we found changes in 6MWD_M_ at home correlated with changes with both 6MWD_D_ (r=0.33, p=0.02) and 6MWD_M_ (r=0.31, p=0.04) on 30ft walking space. Changes in CE_L_ on 90ft and 30ft correlated with changes in CE_L_ at home (r=0.73, p<0.0001 and r=0.61, p=0.0001). Changes in CE_D_ on 90ft and 30ft correlated with changes in CE_M_ at home, (r=0.47, p=0.002 and r=0.62, p=0.00002). There was similar finding with changes in CE_M_ on 90ft and 30ft changes with CE_M_ at home (r=0.50, p=0.001 and r=0.62, p<0.00001).

## Discussion

This two-center study demonstrates that unsupervised home 6MWT, when combined with heart rate monitoring and conducted on a modified walking course, is safe, feasible, and associated with high participant compliance across a broad range of individuals with pulmonary hypertension and heart failure. A cohort of non-cardiopulmonary limited controls was included to explore and establish a benchmark for highly functional data in the home 6-minute walk setting, allowing us to better understand the performance characteristics of the metric in a less-impaired population. We found that sensor-derived CE and 6MWD estimates, using two distinct methods, closely matched directly observed values collected in the clinic, supporting their validity for remote monitoring. As expected, all three measurement methods (direct observation, lap estimation, MAD) showed that 6MWD was greater (~11%) on a 90-ft walking course compared to a 30-ft course or the unsupervised home setting. This difference is likely due to the reduced number of turns required on the longer course. Incorporating heart rate monitoring during the 6MWT and calculating CE helped account for variable effort in the home, providing measurements similar to those observed in the clinic. Changes in CE measured at home were strongly associated with changes observed in the clinic, highlighting the utility of this monitoring approach for tracking changes in physiologic or functional status remotely. This novel method offers broad applicability, particularly in rural, urban, or lower socioeconomic settings where clinic access may be limited, as well as in specialized care centers where patients frequently travel long distances for routine follow-up or assess for treatment response.

There is interest with incorporating passive monitoring using wearable devices in pulmonary hypertension and heart failure. Actigraphy has been included as endpoints in randomized controlled PAH (18, 19), interstitial lung disease (20–22), and heart failure (23) clinical trials. A common finding to all studies has been low activity time; likely from behavioral changes, deconditioning, and underlying cardiopulmonary disease. This makes assessing for therapeutic changes difficult. In PAH, top bursts of activity, peak 5-minute steps, has been shown to increase after adding therapy in two uncontrolled studies(24, 25). More recently, heart rate has been incorporated into the analysis of daily steps to provide more granular insights into the physiologic changes involved in achieving steps (26). Peak steps 5-minute steps and Daily Heart Rate per Step are passive measurements of unstructured home 6MWT and CE. Instead of relying on wrist-based step classification algorithms, that have biases, and error in heart rate measurement with photoplethysmography(3, 4, 6), a structured home 6MWT with a chest-based sensor seems easier to perform, analyze, and trend over time.

There are multiple ways to estimate 6MWD at home including wearables(6, 27), smartphones (28, 29), or global position satellite(30). The type of method can influence the results depending on where it is performed. When using a smartphone app to estimate 6MWD in pulmonary hypertension(31), there was an average difference (standard deviation) of 15 m (76) from the same walk when comparing app estimated 6MWD to directly observed 6MWD. In contrast, when 6MWT was completed outside, presumably with less turns, and compared to directly observed 6MWD indoors, there was an average difference of 2 m (47). These differences highlight the impact the type of monitoring can have when estimating 6MWD and where they can best be implemented. We showed two easy methods for estimating 6MWD based on changes in accelerometry (mean amplitude deviation) and counting laps as long as the course distance is known(6). Measuring truncal movement through a chest-based sensor decreases artifact and allows for estimates to be made in an indoor setting. There was high correlation with directly observed 6MWD and lap estimated 6MWD on 30 ft and 90 ft walking courses. We did find this approach had more error in lower 6MWD when a participant may stop and rest. This can interfere with detecting turns. In this instance, changes in MAD could help correct or verify walk distance. The added benefit of 6MWD_M_ is course distance is not needed for estimation. As long as a walking gait is unchanged during monitoring, this value could be easily trended over time. Not only can 6MWD_M_ be performed at home, it could also serve in real time to verify 6MWD in a clinical trial to make sure there are no large walk discrepancies due to counting errors. In this present study, we found one error in data entry after identifying a discrepancy between directly observed 6MWD with sensor estimated 6MWD. The source document was reviewed and the reported 6MWD matched with the sensor estimated 6MWD.

By incorporating continuous heart rate monitoring during the 6MWT and calculating CE, we can gain insight into the physiologic stress needed to achieve a certain 6MWD. CE has been shown previously to have less variability than 6MWD, correlates strongly with stroke volume, and tracks with clinical improvement (as cardiac output improves the number of heart beats used during 6MWD decreases)(3–6). Especially in an unsupervised home setting, CE helps provide context for a given estimated 6MWD, specifically whether changes were due to worsening physiology or effort. In this report, we found the greatest value of CE was the changes observed in clinic correlated very strongly with changes at home. This provides an opportunity to monitor therapeutic titration or higher risk patients more closely with sensors.

In the initial reporting of a home 6MWT in PAH (10), participants completed in-clinic and home 6MWT on 90ft walking space with a study team member supervising the walks and found similar 6MWD. Having access to a 90ft walking space year-round with supervision by a healthcare team member at home is not a realistic way to complete testing. In our initial report (6) we found unsupervised estimated 6MWD at home on a modified walking space was lower than directly supervised 6MWD on a 90 ft walking space. At the time, we were unsure if this was due to effort or extra turns. By comparing 90 ft to 30 ft 6MWD in clinic on the same day, we showed the course length (and resulting turns) will influence home 6MWD independent of effort when not supervised, and highlights the value of CE. A follow up study used an app to perform tele-6MWT in a mix group of pulmonary hypertension patients and found tele-6MWT had a bias of 25 m over directly observed 6MWT (11). Unless it is related to learning the test, it is hard to image how someone would walk further when unsupervised at home compared to directly supervised 6MWT. Future studies are needed to compare the best mode of estimating 6MWD.

Unsupervised home 6MWT is safe. Including this study, there have now been five reports showing unsupervised home 6MWT can be completed without tripping on oxygen tubing, syncope, or emergency room visits (6, 10, 11, 31). Feedback from participants during this study included a discomfort of doing the 6MWT outside, a similar finding that has been shown previously(11, 30). The indoor test provides comfort about being able to stop and rest if needed, not having to worry about bringing oxygen outside, weather, or walking back once completed. Participants also reported that they liked doing the test in their home as compared to the clinic and got them moving more at home. Therefore, having a home indoor option is extremely valuable for acceptance.

There are limitations to our study. Participants were enrolled as they were seen in the clinic and therefore clinical status could change as there was no screening period for stability. The study was completed during the COVID-19 pandemic and infections impacted follow up. Because of COVID related restrictions with scheduling, there was no room to schedule follow up outside of the week 8 window. We did not design this study to assess therapeutic response. There was electrocardiography data loss when using the manufacturer recommended body location on some patients. A dry electrode chest strap can be an alternative to patch sensors for Cardiac Effort. A potential limitation is the learning effect associated with the unsupervised nature of the at-home version of the test.

In conclusion, in this two-center study we confirm the previous finding that wearable devices with electrocardiography and accelerometry provide a way for 6MWT and CE to be performed safely in the home setting in pulmonary hypertension and heart failure. The measurements track in the same direction as clinic assessments overtime. More frequent assessments between visits could allow for changes in physiology to be identified sooner. Home 6MWT and CE could be performed in all settings (rural or urban) at a fraction of the cost of other tests. Importantly, there was high patient acceptance and compliance with testing. Future studies can compare the effectiveness of using home 6MWT and CE to help with risk assessment or therapeutic response.

## Acknowledgements

Medidata Solutions (formerly MC10, Inc) was instrumental in designing the sensor study and supporting study execution with their expertise of the BioStamp nPoint platform and knowledge of the use of wearable sensors. We would like to thank the clinical operations support of Maura Buckley and Ellora Sen-Gupta (Medidata) for their study support and Paolo DePetrillo and James Caccese (Medidata) for data management, quality control, and processing.

## Data Availability

All data produced in the present study are available upon reasonable request to the authors

